# HFE genotype influences substantia nigra iron accumulation and Parkinson’s disease progression

**DOI:** 10.64898/2026.06.02.26354750

**Authors:** Guangwei Du, Ernest Wang, Christopher Sica, Rachael Stone, Sol De Jesus, Lan Kong, Richard B. Mailman, Xuemei Huang

## Abstract

Increased iron in the substantia nigra has been thought to be a mechanism potentially related to the etiology and/or progression of Parkinson’s disease (PD). We hypothesized that genetic variants of *HFE*, a major iron regulatory gene, would influence substantia nigra iron accumulation in PD. The *HFE* genotype was obtained from 195 subjects (102 PD and 83 Controls) who participated in the PD biomarker program (PDBP) in central Pennsylvania, United States. For this study, carriers of two SNPs (*HFE* H63D and/or C282Y) were considered as variants and the others as wildtype. Susceptibility MRI metrics (QSM, R2*) were assessed at baseline, 18, and 36 months. The primary region of interest was the substantia nigra, the key pathology focus of PD. Group differences in substantia nigra QSM and R2* between HFE variants carriers and wildtype were compared between PD patients and controls at baseline and in progression over time using linear mixed-effects model. We also used interaction analyses to explore if *HFE* genotype impacts clinical measures of PD progression. Of the 102 PD patients, 72 were wildtype, and 30 *HFE* variant. Of the 83 controls, 56 were wildtype and 27 were *HFE* variants. There was a total of 451 data points available for analysis. Compared to wildtype patients, patients with *HFE* variants showed higher baseline substantia nigra QSM (p=0.006), but not higher R2* (p=0.487). Controls had no HFE-dependent differences. Longitudinally, substantia nigra QSM and R2* increased significantly over both 18- and 36-months regardless of *HFE* status (p’s<0.05). Compared to wildtype, PD subjects with *HFE* variants showed an overall faster increase in R2* (p=0.004) and QSM (p=0.003) over the total 36-month epoch, and this reached the statistical significance for R2* during the first 18-months (p=0.026) and for QSM in 36-months (p=0.005). *HFE* status showed a significant interaction with motor scales [MDS-UPDRS II (p=0.006), III (p=0.0002)], suggesting a faster symptomatic progression in PD patients with *HFE* variants compared to wildtype. Although *HFE* genotype has been shown not to associate with the occurrence of PD, these data demonstrate for the first time that in PD patients substantia nigra iron accumulation and disease progression are affected by *HFE* genotype. The underlying mechanisms may be important in the progression of PD and the development of personalized treatment.

## Introduction

Parkinson’s disease (PD) is marked pathologically by dopamine neuron loss in the substantia nigra pars compacta.^1,2^ Postmortem studies have shown higher substantia nigra iron concentrations in PD patients than in controls,^3-6^ but the exact cause of substantia nigra iron accumulation and its role in PD is currently unknown. Excess intracellular iron has been associated with oxidative stress, cellular dysfunction, and neuronal death,^7-10^ thus substantia nigra iron accumulation is hypothesized to play a role in PD pathogenesis^10-12^ and may be a biomarker for PD progression.^13,14^ One clinical trial, however, using the iron chelator deferiprone, found that the drug decreased iron content in nigrostriatal structures, but worsened motor signs in newly diagnosed PD patients over a period of 36 months.^15^ Although disappointing for its intended outcome, the results suggest that iron dysregulation/modulation, particularly in the substantia nigra, may affect PD progression.

These data raised the question of how mechanisms that affect iron metabolism (e.g., regulation of import and export of intracellular iron^10,16^) affect the increased substantia nigra iron accumulation seen in PD. One of these mechanisms is the human homeostatic iron regulator protein (HFE) encoded by the *HFE* gene.^17,18^ HFE regulates cellular iron import directly by competitively binding to transferrin receptor 1 (TFR1) and, indirectly, by affecting iron export via modulation of levels of hepcidin via the iron exporter ferroportin. In preclinical studies, the common *HFE* variant H63D results in partial loss of HFE binding capacity to TFR1, while the less common variant C282Y results in complete loss of HFE expression at the cell surface.^19^ Both variants lead to increased bioavailability of ingested iron by increasing net transport of iron across the intestinal epithelium, and are hypothesized also to influence iron transport across the blood brain barrier.^20,21^ This led to the hypothesis that *HFE* variants (H63D and/or C282Y) may be etiological factors for PD, even though gene association studies have not found an association with increased PD occurence.^22-25^ There are, however, no studies that have focused on testing if *HFE* variants affect PD progression, in part due to a lack of quantitative objective markers for progression.

Recent advances in MRI technology provide a robust way to quantify brain iron *in vivo*.^14,26,27^ Susceptibility MRI is particularly suitable for this task, and its values correlate well with post-mortem chemical measurements of iron in deep gray matter regions.^28,29^ We and others have used both apparent transverse relaxation rate (R2*) and newer quantitative susceptibility mapping (QSM) to estimate iron in the substantia nigra, the key pathological foci of PD.^13,26,30^ The QSM technique commonly used, however, has a significant limitation. There is an arbitrary constant shift in the obtained QSM values due to singularity of dipole kernel at the k-space center.^31^ This can be extremely problematic when using QSM longitudinally. For correction, a zero-referencing technique has been devised that subtracts a mean QSM value from the lateral ventricle area with zero susceptibility where the cerebrospinal fluid is the main source of signal.^31^ Most recently, we demonstrated that the zero-referencing QSM of the substantia nigra can track PD progression over a one-year epoch.^32^ We now use this approach to investigate the effects of *HFE* genotype on nigral iron concentration both cross-sectionally and longitudinally to test two hypotheses: 1) *HFE* genotype affects nigral iron deposition in both PD patients and control subjects at baseline; and 2) *HFE* genotype impacts nigral iron progression measured by QSM and R2*. In addition, we explored if *HFE* genotype affected the clinical measures of PD progression.

## Methods

### Study design and subjects

One hundred-and-two PD patients were recruited from a tertiary movement disorders clinic, and 83 controls were recruited from spouses and the local community in central Pennsylvania as part of the PD Biomarker Program sponsored by the National Institute of Neurological Disorders and Stroke. PD diagnosis was confirmed based on the Movement Disorder Society criteria.^33^ All participants gave written informed consent in accordance with the Declaration of Helsinki, and the protocol was approved by the Penn State Hershey IRB.

All participants were free of major medical issues or neurological conditions other than PD. Demographic and clinical characteristics including the Movement Disorder Society Unified PD Rating Scale parts I, II, and III (MDS-UPDRS-I, -II, -III); levodopa-equivalent daily dosage (LEDD); the Hamilton Anxiety Rating Scale (HARS); the Hamilton Depression Rating Scale (HDRS); and the Montreal Cognitive Assessment (MoCA) were obtained from all participants while patients were on optimized antiparkinsonian medications (ON state) by a movement disorder specialist. Disease duration for PD patients was defined based on the time since first documented PD diagnosis by a physician.

Detailed demographic data from baseline are provided in Table 1. All participants were followed up at 18 months and 36 months to obtain both clinical and MRI data. Eight-two PD patients and 65 controls came back for the 18-month visit, and sixty-three PD patients and 56 controls came back for the 36-month visit. There was no significant difference between dropouts and remaining participants in terms of demographics or *HFE* status.

**Table 1.** Demographic and clinical measures at baseline.

|  | Control (n=83) |  |  | PD (n=102) |  |  |
| --- | --- | --- | --- | --- | --- | --- |
|  | WT (56) | HFE (27) | P-values | WT (72) | HFE (30) | P-values |
| **N at each visit (Baseline/18m/36m)** | 56/43/36 | 27/22/20 | - | 72/59/47 | 30/23/17 | - |
| **Sex, F/M** | 34/22 | 9/18 | 0.034 | 36/38 | 14/16 | 1.00 |
| **Age, years** | 66.7 (9.7) | 66.1 (11.3) | 0.815 | 67.6 (9.2) | 64.9 (9.4) | 0.210 |
| **Disease Duration, years** | - | - | - | 6.2 (6.4) | 6.9 (5.2) | 0.372 |
| **LEDD** | - | - | - | 658 (475) | 776 (463) | 0.255 |
| **MDS-UPDRS I** | - | - | - | 12.3 (7.9) | 9.0 (5.3) | 0.082 |
| **MDS-UPDRS II** | - | - | - | 11.3 (10.3) | 9.1 (9.6) | 0.617 |
| **MDS-UPDRS III** | - | - | - | 29.8 (20.1) | 28.8 (20.4) | 0.354 |
| **HAMA** | 25.9 (2.6) | 24.6 (2.3) | 0.208 | 24.4 (3.0) | 24.2 (4.4) | 0.528 |
| **HAMD** | 4.7 (5.0) | 3.3 (3.2) | 0.495 | 7.9 (6.8) | 6.2 (4.3) | 0.291 |
| **MoCA** | 25.7 (2.6) | 25.0 (2.0) | 0.570 | 23.6 (3.8) | 24.3 (4.4) | 0.830 |

Values for sex are numbers of subjects. Values for age and all clinical measures are means and standard deviations in parenthesis. The p-value for sex is from the Fisher's exact test, the p-values for age, disease duration, and LEDD are from the two-tail Student t test, and p-values for other clinical measures are from ANCOVA with adjustment for age and sex.
Abbreviations: FoG: freezing of gait; LEDD: levodopa equivalent daily dosage; MDS-UPDRS: Movement Disorder Society-Unified Parkinson's Disease Rating Scale; MoCA: Montreal Cognitive Assessment; HAMA: Hamilton Anxiety Rating Scale; HAMD: Hamilton Depression Rating Scale; SD: Standard Deviation.

### *HFE* genotyping

Following an 8-12 hour overnight fast, blood samples were collected from participants by at 10:00h using BD Vacutainer™ glass blood collection tubes with K_3_EDTA (lavender top; Fisher catalog #02-685-2B). Samples were centrifuged at 1,700 x G for 10 min. The supernatant first was aliquoted into cryovials (1 mL each), and then the buffy coat was removed gently and also placed in a cryovial. The samples were stored at -80 C. The buffy coats were used to extract DNA for HFE genotyping for each subject using polymerase chain reaction-restriction fragment length polymorphism analysis, as described previously.^34^ To simplify writing, when referring to individuals without either of the *HFE* H63-C282 genotypes we use the term <u>wildtype</u>. Conversely, those individuals with H63D and/or C282Y are referred to as <u>variant</u>. Of all participants, in the wildtype and variant groups, there were two homozygous H63D carriers, one homozygous C282Y carrier, and two compound heterozygous carriers. They were included in the study and categorized as HFE variant carriers.

### MRI Image acquisition and analysis

For each participant, T1-weighted, T2-weighted, and multi-gradient-echo MR images were acquired on a 3T Siemens scanner (Erlangen, Germany). A magnetization-prepared rapid acquisition gradient echo sequence was used to obtain T1-weighted images with repetition time/echo time = 1540/2.34 milliseconds; field of view = 256×256; slice thickness, 1 mm (with no gap); and slice number = 176. A 3-dimensional T2-weighted SPACE (sampling perfection with application optimized contrast using different angle evolution) sequence was used to obtain T2-weighted images with repetition time/echo time = 2500/316 milliseconds and the same spatial resolution settings as the T1-weighted images. T2*-weighted images were acquired using a multi-gradient-echo sequence with 8 echoes (echo times evenly spaced from 6.2 to 49.6 milliseconds); and repetition time = 55 milliseconds; flip angle = 15°; field of view = 240×240; matrix = 256×256; slice thickness = 2 mm; slice number = 64; and voxel size = 0.9×0.9×2 mm^3^. All images were inspected offline and deemed free of severe artifacts or any major structural abnormalities. Quantitative susceptibility maps (QSM) were generated using morphology enabled dipole inversion{Liu, 2012 #2654} using an automatic uniform cerebrospinal fluid zero reference (MEDI+0) with a nonlinear formation of the magnetic field to source.^31^

The substantia nigra region was segmented using automatic atlas-based parcellation, followed by manual correction.^14^ This semiautomatic approach was designed to improve the accuracy and repeatability of the segmentation and consisted of four steps. First, T1- and T2-weighted images from all participants were used to construct a cohort-specific template using an unbiased atlas construction algorithm in the Advanced Normalization Tools (ANTs) package.^35,36^ Since most of these structures have high iron content and are seen best in T2-weighted images, the ROIs were defined manually on the constructed T2-weighted template according to previous studies.^14,37^ Figure 1 shows the exact location of these six structures. Second, an atlas-based segmentation pipeline (AutoSeg v3.0; University of North Carolina Neuro Image Analysis Laboratory) generated the substantia nigra segmentation in individual space based on the T2-weighted images. Third, automatic segmentation results from all subjects were inspected visually and corrected manually by a rater blinded to group information. Lastly, an affine registration algorithm was used to bring the ROIs from T2-weighted images to the QSM and R2* images by registering to an averaged magnitude image. Finally, mean QSM and R2* values from both sides were calculated for individual subjects.

**Figure 1.**
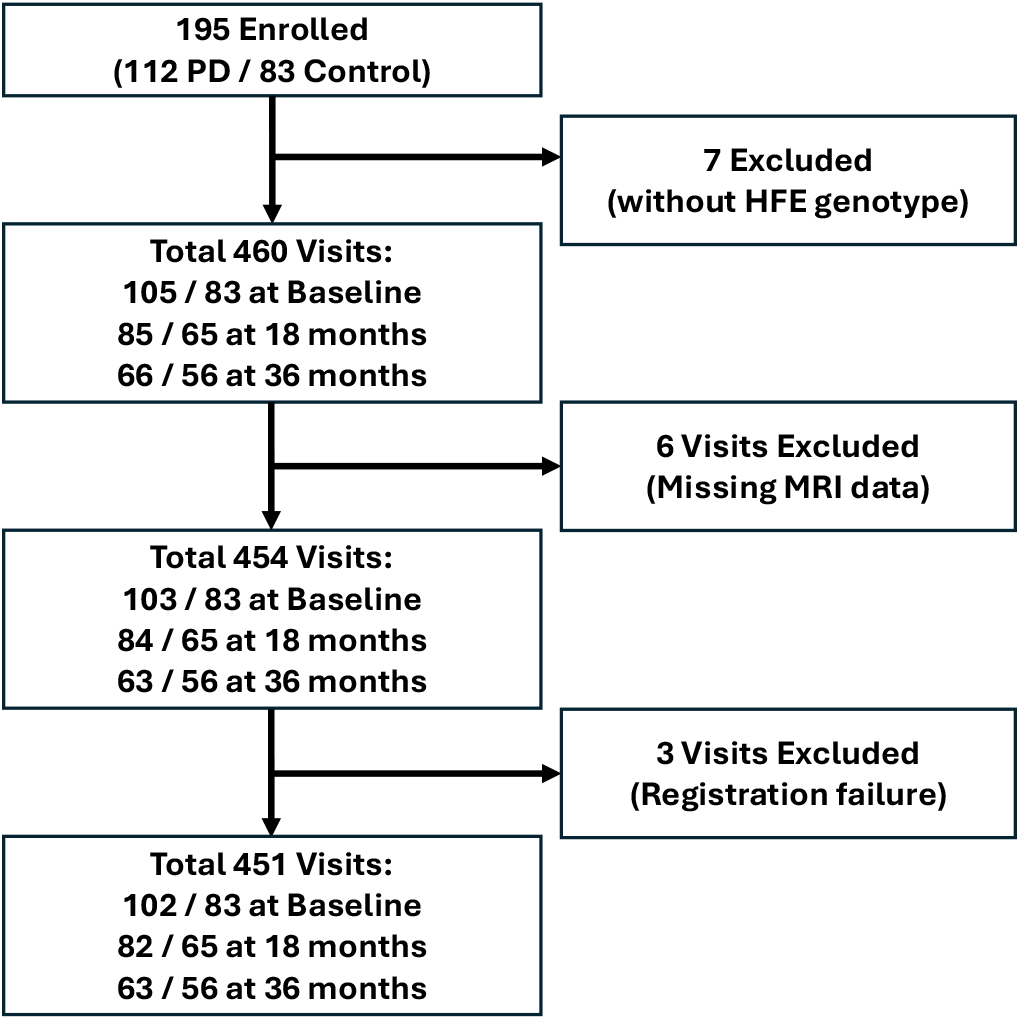
Flowchart for sample size determination.

### Statistical analysis

#### Group comparisons of demographics and clinical measures at baseline

*HFE* variants frequencies were compared between PD patients and controls using Fisher’s exact test. Demographic data were compared between subjects with and without *HFE* variants using Fisher’s exact test for sex and Student’s *t-*test for age separately for PD patients and controls. Disease duration and LEDD were compared between patients with and without *HFE* variants using one-way analyses-of-covariance (ANCOVA) adjustment for age and sex. Clinical measures (MDS-UPDRS-I, -II, and -III MoCA, HAMA, HAMD) were compared between subjects with and without HFE variants using ANCOVA with adjustment for age, sex, disease duration and LEDD for PD patients and adjustment for age and sex for controls.

#### Group and longitudinal analyses of MRI data

A two-way ANCOVA with the Disease Group × *HFE* status interaction term, and age and sex as covariates was used to compare substantia nigra QSM and R2* values at baseline. Post-hoc tests were used to evaluate the effects of *HFE* variants separately for PD patients and controls. The Tukey-Kramer method considering six QSM or R2* pair-wise comparison was used for multiple comparison correction in group comparisons of regional MRI values at baseline. Linear mixed models were used for longitudinal analysis of progression of substantia nigra QSM and R2* values with baseline age and sex as covariates and intercept and visit time as random variables. Bonferroni correction was used to control multiple comparison by a factor of 2 (two MRI measures, significant if p < 0.025).

#### The impact of *HFE* on clinical measures

Linear mixed-effects models were used to test whether HFE has impact on group difference and progression of clinical measures within 36 months epoch with adjustments of age, sex, disease duration, and LEDD at baseline. To further explore the impact of *HFE* gene between clinical measures and disease duration across whole study cohort, we used linear regression models with clinical measures as response variables, and Disease Duration, *HFE* status, and Disease Duration × *HFE* status as predictors, age, sex, and LEDD also are included in the model as covariates. The significant interaction term in the above-mentioned model indicates that *HFE* has impact on disease progression within the study cohort. All statistical analyses were performed using SAS 9.4 M9 (SAS Institute Inc., Cary, NC).

## Results

### Study flow and demographics

The overall design enrolled 195 subjects (Figure 1). Seven subjects were excluded from analyses due to the lack of HFE genotype information, six visits were excluded due to missing MRI data, and three patient-visits were excluded due to image registration failure. Eight-two PD patients and 65 controls returned for the 18-month visit, and 63 PD patients and 56 controls returned for the 36-month visit. The final analysis included 102 PD patients (72 wildtype, 30 variants) and 83 controls (56 wildtype, 27 variants) with a total of 451 data points available for analysis (see details in Figure 1).

The analysis of the demographic and clinical data for each group (Table 1) found no significant difference between PD patients and controls in *HFE* variant frequencies (p = 0.734). There also were no significant differences between subjects with or without *HFE* variants in terms of gender, age, and other clinical measures in PD except that PD patients with *HFE* variants showed a weak trend for lower baseline MDS-UPDRS I (p = 0.082) scores. In controls, there were more males with *HFE* variants (male:female ratio = 1:2).

### Influence of HFE genotype on nigral iron accumulation in cross-sectional analyses

At baseline, there were significantly higher substantia nigra QSM and R2* values in PD patients compared to controls. Within PD, patients with *HFE* variants showed higher substantia nigra QSM values compared to the wildtypes (corrected p=0.004; see Figure 2). Within controls, there were no significant differences between *HFE* wildtype and variant carriers in the substantia nigra QSM or R2* (corrected p = 0.999). There was no difference in substantia nigra R2* between *HFE* wildtype and variants among PD patients or controls.

**Figure 2.**
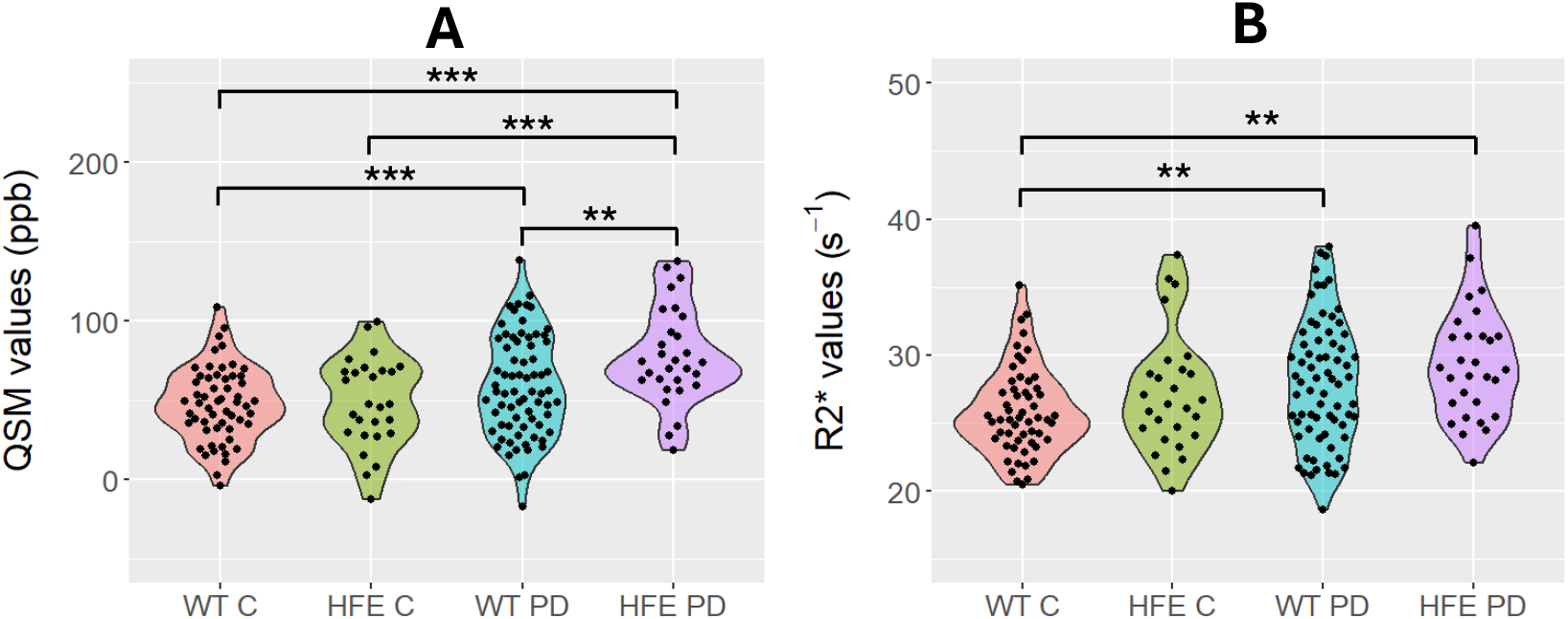
Baseline group comparisons of nigral iron levels measured by QSM (panel A) and R2* (panel B). * Indicates p < 0.05; ** indicates p < 0.01, *** indicates p < 0.001. p values are after Tukey-Kramer correction. WT = wildtype HFE subjects, HFE = subjects with HFE variants, C = healthy controls, PD = Parkinson’s patients.

### The impact of HFE on nigral iron progression in longitudinal analyses

Compared to controls, both substantia nigra QSM and R2* increased significantly over 36 months indicated by significant p-values for Time effect (Figure 3 and Table 2). The PD patients with HFE variants had a faster substantia nigra QSM progression compared to wildtypes [Time × Group effect: F(1,143) = 8.96, p = 0.0033] over the total 36 months. Patients with *HFE* variants also showed a faster substantia nigra R2* progression than did patients with WT [F(1,143) = 8.33, p = 0.0045] over the total 36 months. Compared to PD with HFE wildtype, HFE variants showed a faster progression in substantia nigra QSM over the second 18-months (F(1,58) = 8.39, p=0.0053), whereas in substantia nigra R2* during the first 18-months [F(1,79) = 4.82, p=0.026].

**Table 2.** Longitudinal analysis of SNc QSM and R2* from linear mixed models.

| | | Time Effect | Group Effect | Time × Group |
| --- | --- | --- | --- | --- |
| SNc QSM | 0-18 mo | **0.0005** | **0.0172** | 0.1558 |
|  | 18-36 mo | **0.0025** | 0.3455 | **0.0053** |
|  | 0-36 mo | **<0.0001** | 0.0391 | **0.0033** |
| SNc R2* | 0-18 mo | **0.0249** | 0.5622 | 0.0261 |
|  | 18-36 mo | **0.0004** | 0.1173 | 0.5778 |
|  | 0-36 mo | **<0.0001** | 0.3426 | **0.0045** |

The values are $p$ values of variables of interest from linear mixed models. Bold indicates significance after Bonferroni correction.

**Figure 3.**
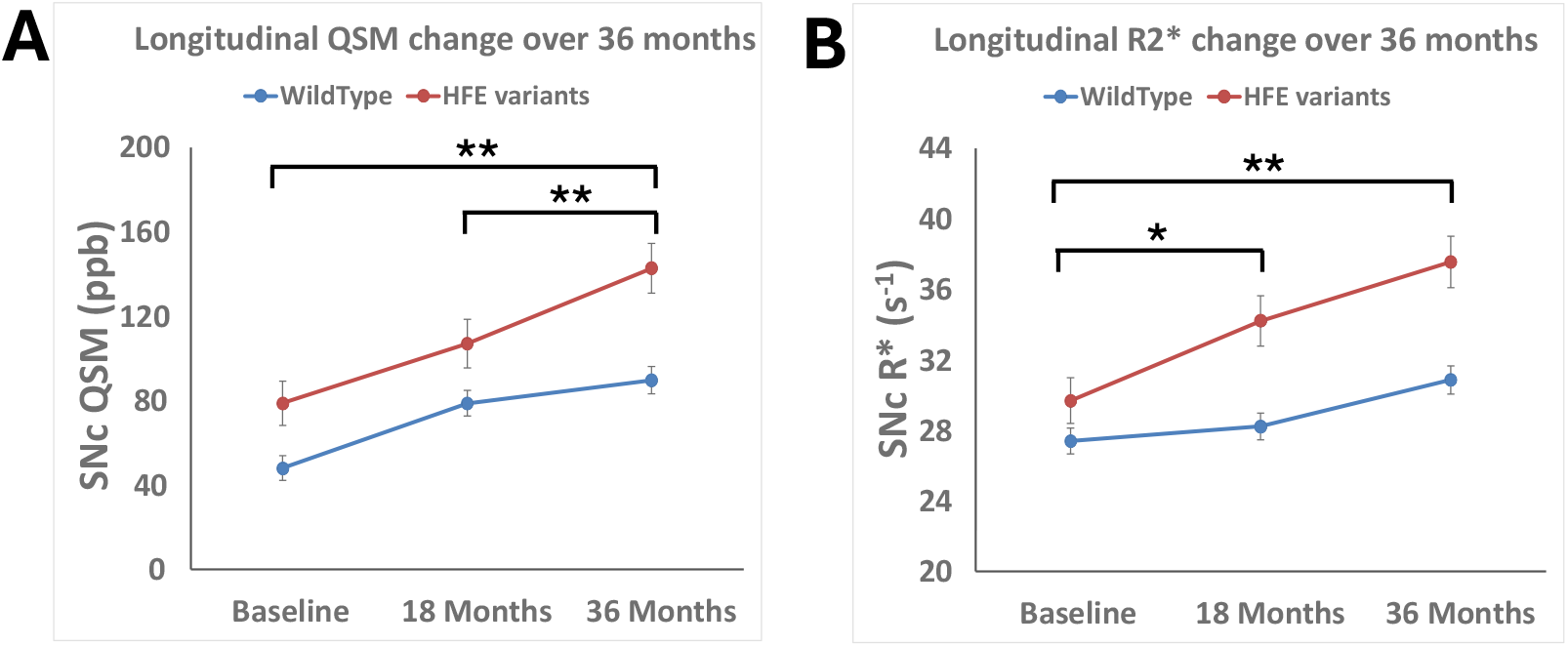
Longitudinal effects of HFE on SNc QSM and R2* values over 36 months. Error bars are SEM. Means and SEM for each time points were estimated using linear mixed models with baseline age and sex as covariates. * Indicates p < 0.05; ** indicates p < 0.01.

### The impact of HFE on clinical progression

The linear mixed-effects models were used to explore group difference and longitudinal progression of each clinical measure independently (Table 3). There were no significant differences between patients with and without *HFE* variants in terms of clinical measures or clinical progression over the 36-month epoch. The interaction terms between *HFE* genes and disease duration from regression analyses, however, were significant and suggested that *HFE* status had a significant modifying effect on clinical progression. Namely, the data over a longer period of timeframe (as much as 20 years) showed that PD patients with the HFE variants displayed a faster clinical progression than wildtype patients (Figure 4).

**Table 3.**
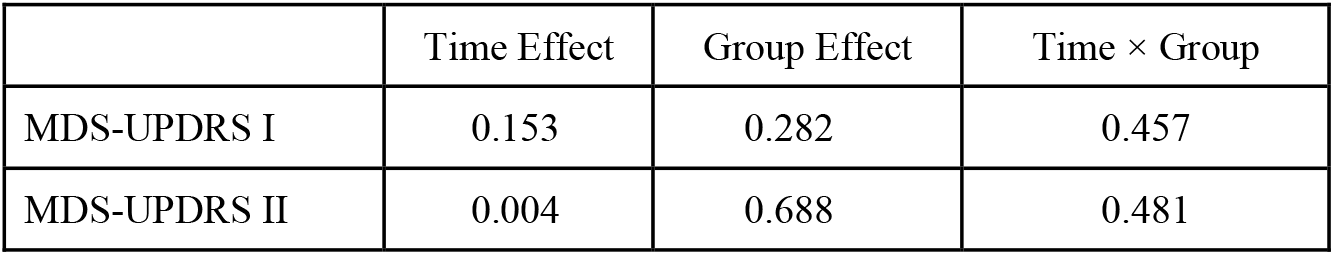
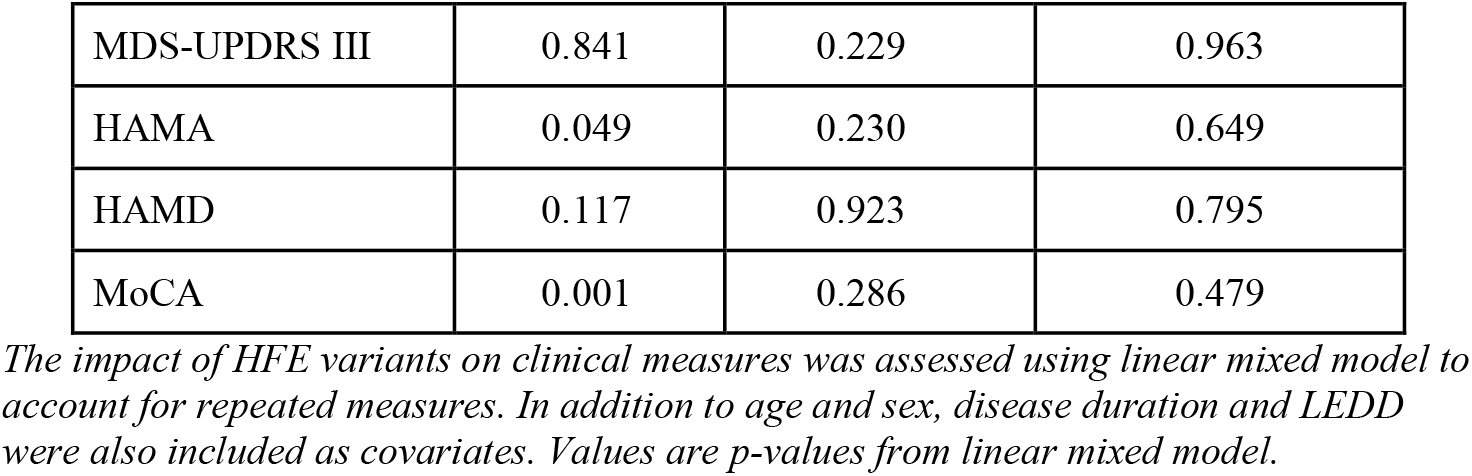
Direct HFE impact on clinical measures using linear mixed-effect models.

**Figure 4.**
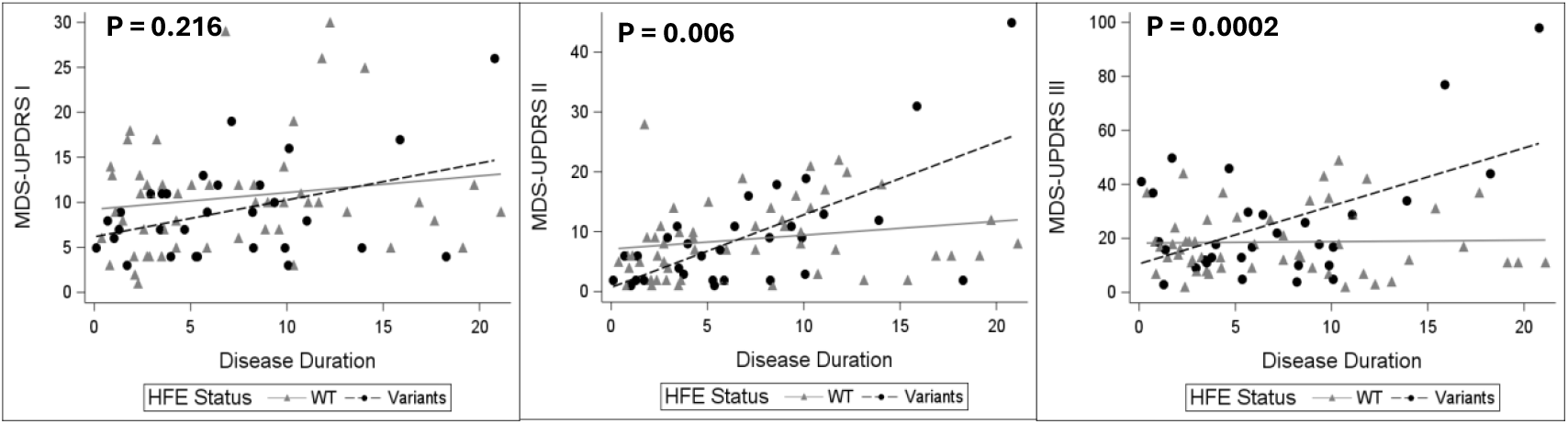
HFE moderates the associations between clinical measures and disease duration.

## Discussion

These data show for the first time that common *HFE* variants (H36D and C282Y) have a significant impact on nigral iron content in Parkinson’s patients. In those without clinical diagnoses of PD, however, the *HFE* genotype did not affect nigral iron content. Using state-of-art zero-referencing QSM, we also showed that PD patients with *HFE* variants had a faster nigral iron accumulation over 36 months than wildtype PD patients. In addition, we discovered that *HFE* variants may predict faster progression of motor symptoms over ∼20 years. In total, this study supports the unique role of nigral iron dysregulation in PD progression and how *HFE* variants may represent a genotype that helps predict PD progression in terms of both substantia nigra iron accumulation and clinical outcomes. This warrants further investigation to understand the mediating mechanisms and the potential implication for individualizing treatment strategies in PD.

### The distinct roles of HFE gene variants in individuals with and without PD

*HFE* variants are known to result in loss-of-function of HFE proteins, which in turn, can leads to the increased brain cellular iron accumulation.^21^ Although higher cellular iron content may be beneficial in healthy athletes,^38^ it can also increase oxidative stress by forming neurotoxic redox couples. Indeed, it has been hypothesized as a contributing mechanism in neurodegeneration such as Alzheimer’s disease and amyotrophic lateral sclerosis.^10,16^ As noted earlier, numerous studies have examined potential links between *HFE* variants and PD occurrence, but no convincing relationships were found.^22-24,39^

Advances in MRI technology, such as QSM, have offered a powerful means to study tissue iron in vivo, and they are being used to examine brain tissue iron in PD patients.^13,14,26^ There was, however, no prior study to investigate if *HFE* gene status affects nigral iron content in Parkinson’s disease. Our study provides the first evidence that *HFE* status has a significant impact on nigral iron content in PD. This was captured by baseline QSM metrics that are more specific for local iron measurement. R2*, another susceptibility MRI metric, also trended higher in *HFE* variants, although it did not reach statistical significance. This difference may be due to the fact that R2* not only captures iron-related changes, but also local cellular pathological changes in the substantia nigra due to PD-related processes. Future studies including other microstructural-specific metrics may help resolve this.

There are two published studies that examined the impact of *HFE* on brain iron content. Kalpouzos et al.^40^ studied a cohort of 202 healthy subjects, 66 *HFE* variants and 142 wildtypes. They reported that those with *HFE* variants have a higher load of iron in the putamen and that it was associated with higher executive function. The study did not include the substantia nigra in their analyses. Bartzokis et al.^41^ studied 35 male healthy subjects, 20 with iron regulatory gene variants (H63D and transferrin C2 allele) and 15 wildtypes, using field-dependent relaxation increase (FDRI), and an older susceptibly MRI technique to estimate iron. They reported a higher FDRI value, suggesting higher iron in the caudate nucleus in male iron gene variant carriers. They also studied 31 females (15 variants and 16 wildtypes) but reported no *HFE* differences.^41^ Thus,the current study is the first to focus on the substantia nigra in patients with PD and healthy subjects.

Our finding suggests that *HFE* gene status itself does not impact iron accumulation in healthy subjects. This may provide an explanation why these *HFE* variants alone are not risk factors for developing PD. The finding of robust substantia nigra iron accumulation only after PD diagnoses is consistent with the possibility that a PD treatment effect (e.g., levodopa) may be involved.^42^ This hypothesis may be part of reason why the clinical trial with the iron chelator deferiprone failed,^15^ as nigral iron accumulation was not high enough to be detrimental to PD patients who were drug naïve.

### The impact of HFE genotype in substantia nigra iron progression

Longitudinal progression of nigral iron accumulation was not possible until susceptibility MRI was available.^14,43,44^ In 2018, our group reported the first longitudinal changes in nigral iron accumulation in PD patients using R2*.^26^ We did not detect significant longitudinal changes using traditional QSM.^14^ In the current study, we used CSF-zero-referencing to provides a more precise QSM estimation without an arbitrary shift of zero values during longitudinal examination. We recently reported that the CSF zero-referencing QSM captured a faster substantia nigra iron accumulation (one-year epoch) in PD patients compared to control with a large effect size (Cohen’s *d* = 1.14 vs 0.42 for clinical UPDRS score).^32^ Zero-referencing QSM appears to provide a new lens to study longitudinal changes of substantia nigra iron and the impact of *HFE* gene.

In the current study, we found that PD patients overall have faster substantia nigra iron accumulation captured by both QSM and R2* over 36 months epoch. This is consistent with our previous findings with early-stage PD patients using the same zero-referencing QSM.^32^ The exact mechanisms for faster substantia nigra iron in progression in PD is unknown. It has been postulated that blood brain barrier disruption in neurodegenerative conditions subsequently leads to iron dysregulation.^45^ Our recent discovery of low-to-normal nigra iron in drug-naïve PD patients suggests an intriguing alternative hypothesis; that higher nigral iron accumulation may be a marker of side effect of PD treatment, especially levodopa has been shown to have neurotoxic properties in a number of preclinical studies.^16^ When iron accumulation in the substantia nigra exceeds and overcomes normal homeostatic mechanisms, it may propel ongoing neurodegeneration and clinical progression in patients with PD. The levodopa-toxicity hypothesis is not a new concept and was tested by many previous studies (e.g. ELLDOPA etc.).{Fahn, 2004 #6380} Those well-designed large and costly human studies did not, however, have a sensitive *in vivo* marker for potential levodopa-toxicity. Revisiting this idea, with modern toolset (such as zero-referencing QSM), may transform our understanding of PD pathoetiology and therapeutic approaches.

Heterogeneity of PD progression is well-known but not well understood, and many factors may be involved, including both the patient factors and physician’s choice of treatment strategies. Our finding that *HFE* variants have differential impact on longitudinal nigral iron accumulation in PD may shed light on both factors. Further studies are needed to investigate the interaction effect between *HFE* and levodopa treatment as it may open an exciting possibility to develop individualized monitoring and treatment strategies. For example, one possible future scenario is that *HFE* variants carriers should be monitored closely for their nigral iron content after starting levodopa, and deploying a levodopa-sparing or iron chelating strategy when nigra iron is trending up to cross certain threshold.

### The clinical impact of HFE genotype status on PD patients

Our literature review could find no prior study that had investigated the effects of *HFE* on PD clinical progression in our literature review. In the current study, we did not detect significant differences in the progression of clinical scores over 36 months epoch between *HFE* variants and wildtype in patients with PD. This is not surprising as clinical scores have limited power to detect longitudinal change compared to nigral iron marker as we reported in a recent publication (Cohen’s *d* = 1.14 for QSM vs 0.42 for clinical MDS-UPDRS II score). Regression analysis of clinical measures leveraging disease duration information, however, detected a faster clinical progression over a 20-year timeframe in PD with *HFE* variants. Together, the result from these explorative analyses suggests that *HFE* variants has impact in clinical progression and long-term outcomes and this may be mediated by its impact on nigra iron accumulation. Further studies, with larger dataset and full range of patients’ duration of illness are warranted.

### Limitations

There are several limitations to our study. First, the sample size was relatively small for investigating a genetic variance, although one estimate is that H63D and C282Y are carried by approximately a quarter and a tenth, respectively, of Caucasians in the United States.^46^ Second, although there are technical advantages of using QSM versus R2* for quantifying iron *in vivo*, the QSM signal measures the bulk susceptibility of underlying tissue components that might include contributions from calcium, lipids, or myelin. Moreover, the exact type of iron reflected by QSM is unclear and may include heme or non-heme iron, ferritin- or non-ferritin-bound iron, and does not differentiate iron within neurons versus glia. Third, our MRI and clinical data were captured in the on-drug state, and thus both QSM and clinical measures may be confounded by PD medications. Although we controlled LEDD in our correlation analyses, it is both difficult and impractical to determine the medication effect in mid- and later stages PD patients. Future studies with larger sample sizes (*HFE* variants), preferably on both drug-naïve patients and drug-treated patients, will be essential to better understand the roles of iron and *HFE* variants in PD-related pathophysiology.

In preclinical studies, *HFE* genotypes have been reported to be protective against cellular damage initiated by misfolded α-synuclein via increasing autophagy, thereby increasing elimination of α-synuclein.^47^ In mice, the *HFE* variants genotype also decreased iron in the SN in a mouse model, and provided motor functional benefits.^48^ In our observational human study, however, we found that *HFE* variants accelerated nigral iron accumulation and was associated with faster clinical motor progression. It will be useful to determine why the preclinical and clinical studies appear to diverge, but it underscores the importance of cross-discipline communication and discussion.

## Conclusion

This study was focused on the substantia nigra because it is both the site of major cellular loss in PD, and also where iron accumulation occurs with PD progression. The results, however, cannot be extrapolated to structures outside of the substantia nigra. Future studies should investigate how *HFE* variants influence iron accumulation and disease progression in PD, including the roles of extranigral structures, and specific cell types and pathways involved. Our results also demonstrated that zero-referencing QSM provides a useful marker for future human translation research into this topic.

## Data availability

The data that form the basis of this study are available to qualified investigators upon request.

## Acknowledgements

We express gratitude to all the participants who volunteered for this study and study personnel who contributed to its success.

## Author contributions

1. Research project: A. Conception, B. Organization, C. Execution;
2. Statistical Analysis: A. Design, B. Execution, C. Review and Critique;
3. Manuscript: A. Writing of the first draft, B. Review and Critique;

Guangwei Du: 1A, 1B, 1C, 2A, 2B, 3A.

Ernest Wang: 1B, 1C, 2C, 3B.

Christopher Sica: 1C, 3B.

Sol De Jesus: 2C, 3B.

Rachel Stone: 2C, 2B.

Lan Kong: 2A, 2C, 3B.

Richard Mailman: 1A, 2C, 3B.

Xuemei Huang: 1A, 1B, 1C, 2A, 2C, 3B.

